# Anticoagulation after hemorrhagic transformation in acute cardioembolic ischemic stroke

**DOI:** 10.1101/2024.10.31.24316533

**Authors:** Hyunsoo Kim, Ye-Eun An, Beom-Seok Seo, Jae-Myung Kim, Kang-Ho Choi, Man-Seok Park, Ji Sung Lee, Joon-Tae Kim

## Abstract

**Background:** The safety and efficacy of anticoagulation in the presence of hemorrhagic transformation (HT) in cardioembolic acute ischemic stroke (AIS) remain uncertain.

**Methods:** This retrospective study enrolled patients presenting with cardioembolic AIS within 48 hours at a tertiary stroke center between January 2011 and August 2023. Patients who developed HT during hospitalization and underwent follow-up imaging were included, focusing on those with hemorrhagic infarction or parenchymal hematoma type 1. Primary outcomes were HT exacerbation on follow-up imaging and 3-month modified Rankin Scale (mRS) distribution shift, comparing anticoagulation therapy (AC), antiplatelet therapy (APT), and drug discontinuation (NM). The safety outcome was symptomatic intracerebral hemorrhage (sICH) occurrence.

**Results:** Among 763 patients with HT (mean age 74.6±8.9 years, 48.1% male), AC was associated with a higher incidence of HT exacerbation compared to APT (adjusted OR 0.48, 95% CI 0.29-0.80, p-value =0.005). However, AC demonstrated improved 3-month mRS outcomes versus APT (adjusted OR 0.63, 95% CI 0.43-0.92, p-value =0.017) and NM (adjusted OR 0.38, 95% CI 0.26-0.55, p-value <0.001). sICH occurred in 5% of cases overall, with rates of 1.5%, 2.1%, and 11.7% in the AC, APT, and NM groups, respectively (adjusted OR for NM vs. AC: 3.93, 95% CI 1.18-13.16, p-value =0.026).

**Conclusion:** In cardioembolic AIS patients with HT, excluding those with PH-2, anticoagulation may lead to radiological exacerbation without increasing sICH risk, while potentially improving functional outcomes. These findings suggest that the presence of HT should not necessarily preclude the use of anticoagulation therapy in this patient population.

## Introduction

Hemorrhagic transformation (HT) is considered a natural occurrence in the progression of acute ischemic stroke (AIS) and is often used as an indicator of clinical outcomes in stroke patients.^1^ While certain HT subtypes, such as hemorrhagic infarction (HI) and parenchymal hematoma (PH) type I, may not significantly impact neurological outcomes or mortality, PH type II has been strongly associated with neurological deterioration and increased 3-month mortality rates.^2–4^

HT represents a frequent complication in AIS, with an incidence ranging from 13% to 46% of cases.^5, 6^ Notably, HT tends to manifest with greater frequency and severity in strokes of cardioembolic origin compared to those stemming from other etiologies.^7^

Clinical guidelines are controversial regarding the optimal timing for initiating anticoagulation in cardioembolic AIS.^8, 9^ This presents a significant clinical challenge, particularly regarding the optimal timing of anticoagulation initiation. The decision to initiate anticoagulation therapy in cardioembolic AIS patients involves a delicate balance between mitigating the risk of early recurrent embolic events and potentially exacerbating bleeding complications.^10–12^ Recent large-scale trials have demonstrated that early anticoagulation is non-inferior to delayed initiation in atrial fibrillation (AF) patients with AIS.^13, 14^ However, the presence of HT remains a critical factor in determining the timing of anticoagulation initiation. Despite the significance of this issue, robust evidence concerning the safety of anticoagulation in the context of HT is lacking. The previous existing studies are limited by small patient cohorts or reliance on subgroup analyses, thereby constraining the ability to formulate definitive recommendations regarding anticoagulation after HT.^15, 16^

To address this critical knowledge gap, our study aims to investigate the impact of anticoagulation on HT exacerbation, as visualized on follow-up imaging, and on clinical outcomes in patients with cardioembolic AIS. We have specifically excluded patients with PH-2 from our analysis, given its well-established association with poor neurological outcomes. By focusing on patients with less severe forms of HT, we aim to provide more nuanced insights into the safety and optimal timing of anticoagulation therapy in this population.

## Methods

### Subjects

This retrospective study included patients with acute ischemic stroke who presented within 48 hours of symptom onset at a tertiary stroke center between January 2011 and August 2023 (n=12,658). Among them, image analysis was performed on patients with cardioembolism (CE) classified according to the Trial of Org 10172 in Acute Stroke Treatment (TOAST) criteria (n=2,820).^17^ Only patients who developed HT based on imaging performed during hospitalization were included (n=1,050). Those for whom changes in HT could not be assessed due to a lack of follow-up imaging after the initial HT were excluded (n=896). Additionally, patients with an initial diffusion-weighted magnetic resonance imaging (DWI) Alberta Stroke Program Early CT Score (ASPECTS) score of 0–2, indicating high stroke severity on DWI (n=800), and those with an initial HT type classified as parenchymal hematoma type 2 (PH-2) or higher were excluded, resulting in a final analysis of 763 patients.

### Definition of HT and HT exacerbation

HT was classified radiologically according to the European Cooperative Acute Stroke Study (ECASS) criteria.^3^ Based on the ECASS classification, HT is typically categorized radiologically into two main subtypes: HI and PH. According to ECASS criteria, HI-1 is defined as small petechial hemorrhage within the infarcted area without mass effect, while HI-2 is characterized by confluent petechial hemorrhage within the infarcted area, also without mass effect. PH-1 is defined as a hematoma involving less than 30% of the infarcted area, accompanied by mild mass effect. PH-2, on the other hand, involves hematoma in more than 30% of the infarcted area with definite mass effect. In this hospital, where DWI is extensively used, HT was detected using DWI and susceptibility-weighted imaging (SWI), which offer greater sensitivity in detecting HT compared to computed tomography (CT). Imaging assessments were conducted by a committee of three neurologists specializing in emergency stroke care. Discrepancies in interpretation were resolved through consensus during regular meetings. Exacerbation of HT was defined as an increase in HT severity by one or more stages (e.g., from HI-1 to HI-2) or as an expansion of the hemorrhagic lesion within the same HT type. Symptomatic intracerebral hemorrhage (sICH) was defined as HT of PH-2 or higher, accompanied by worsening neurological symptoms on follow-up imaging.

### Imaging protocol and analysis

The imaging protocol for acute stroke patients at our center is as follows: DWI and magnetic resonance angiography are conducted within a few hours of arrival at the emergency department unless contraindications exist. In more complex cases, CT angiography is performed. When intravenous thrombolysis (IVT) or endovascular treatment (EVT) is administered, DWI is typically conducted 1 day after admission to assess the presence of lesions. Subsequent imaging includes CT angiography on day 3 of hospitalization, if necessary, followed by diffusion MRI on day 5 for final follow-up. Additional tests are performed at the discretion of the attending clinician. Patients included in this study were those whose HT was confirmed through imaging performed within 1 week of hospitalization after the index stroke. Only those patients who underwent follow-up imaging to assess changes in HT were analyzed to evaluate the impact of treatment. Accordingly, we aimed to evaluate the exacerbation of HT. When measuring the ASPECTS score, the focal embolic lesion was excluded from scoring. Only confluent lesions within the scoring region were counted and subsequently analyzed.

Ethnic statements. The current study was approved by local institutional review boards at all participating centers, including Chonnam National University Hospital (CNUH-2024-294).

### Data collection

Demographic, clinical, imaging, and laboratory data were analyzed retrospectively. Imaging tests were conducted at least twice during hospitalization, with the HT type confirmed at each time point, and HT aggravation was assessed according to predefined criteria. The ASPECTS score was measured using DWI performed at the time of initial hospitalization, with separate scores calculated for the anterior circulation and posterior circulation based on previously published studies.^18, 19^ Mean systolic blood pressure (mSBP) was defined as the average of at least six systolic blood pressure measurements within the first 24 hours after admission. Patients were classified according to the treatment method used following the initial confirmation of HT: 1) anticoagulation therapy (AC), 2) antiplatelet therapy (APT), and 3) drug discontinuation (NM). The AC group included patients treated with direct oral anticoagulants (DOAC) or vitamin K antagonists, without distinctions regarding dosage. The APT group consisted of patients who received aspirin, clopidogrel, cilostazol, or ticlopidine, regardless of the type, dose, or combination of drugs. The NM group included patients who discontinued their anticoagulant or antiplatelet therapy or did not resume these medications after HT if they were not already on them. In addition to examining the presence or absence of medication at the time of HT occurrence, antiplatelet and anticoagulation use at discharge was also analyzed. In addition to assessing whether medication was being used at the time of HT occurrence, the study also analyzed the use of antiplatelet and anticoagulation therapy at discharge.

### Outcome

The primary outcomes were the occurrence of HT exacerbation on imaging and the shift in modified Rankin Scale (mRS) distribution at 3 months. Safety outcomes included the occurrence of sICH on follow-up imaging. Secondary outcomes included the proportion of patients with a good mRS (0-2) at 3 months, and the evaluation of composite events at 3 months, such as recurrent stroke (either hemorrhagic or ischemic), myocardial infarction (MI), and all-cause mortality, as well as stroke recurrence.

### Statistical analysis

Baseline characteristics and outcomes were compared according to treatment methods using the chi-square test, ANOVA, Cochran-Mantel-Haenszel shift test, or Kruskal-Wallis test, depending on the type of variable. The presence of HT exacerbation, sICH occurrence, good functional outcomes at 3 months, and vascular outcomes by treatment were analyzed using Kaplan-Meier methods, chi-square tests, and log-rank tests. Odds ratios (ORs) and 95% confidence intervals (95% CIs) were calculated using a logistic regression model to evaluate the impact on HT exacerbation, 3-month mRS, and safety outcomes. Hazard ratios (HRs) and 95% confidence intervals (95% CIs) for 3-month vascular outcomes and stroke recurrence were analyzed using the Cox proportional hazards model for each treatment method. Predefined variables considered clinically relevant were adjusted for: age, male sex, NIHSS score, previous mRS score, ASPECTS, HT type, history of stroke, coronary heart disease, hypertension, diabetes, hyperlipidemia, TIA, peripheral arterial disease, prior diabetes treatment, prior anticoagulation treatment, acute thrombolytic treatment, white blood cell count, total cholesterol, low-density lipoprotein-C, blood urea nitrogen, creatinine, and fasting glucose. Ordinal shifts toward better outcomes were analyzed using common ORs with 95% CIs. Hazard ratios (HRs) for 3-month vascular outcomes and stroke recurrence were assessed for each medication method using a Cox proportional hazard model with the same adjusted variables as in the logistic regression model. Two-sided p-values <0.05 were considered significant. Statistical analyses were conducted using the R software “rms” package (version 3.6.0, R Foundation for Statistical Computing, Vienna, Austria).

## Results

### General characteristics

The average follow-up period for each patient was 86.4 ± 18.5 days. The general characteristics and clinical features of the study population are presented in Table 1. A total of 763 HT patients were included in the analysis (mean age 74.6±8.9 years, 48.1% male). After developing HT, 324 patients (42.4%) received AC, 191 patients (25.0%) received APT, and 248 patients (32.5%) NM. Regarding HT type, 347 patients (45.5%) were classified as HI-1, 322 patients (42.2%) as HI-2, and 94 patients (12.3%) as PH-1. The AC group had a relatively higher proportion of HI-1 cases (n=189, 54.4%) and a lower proportion of PH-1 cases (n=9, 10.0%). In contrast, the NM group had a lower proportion of HI-1 cases (n=61, 17.6%) and a higher proportion of PH-1 cases (n=69, 73.4%), showing a significant difference between treatment groups (p<0.001). The median initial NIHSS score was 11 (IQR 6-15), with a median score of 9 (IQR 3-13) in the AC group and 12 (IQR 8-15) in the NM group (p<0.001). Among the patients, 658 (86.2%) had anterior circulatory infarction, while 105 (13.8%) had posterior circulatory infarction. The ASPECTS score for anterior circulation showed a difference between treatment groups, with a median score of 7 (IQR 6-8) in the AC group compared to 7 (IQR 5-8) in the NM group (p<0.001), while there was no significant difference in the ASPECTS score for posterior circulation. Among the patients who were not on any medication at the time of HT, 29.3% were discharged on anticoagulation, whereas 82.4% of those who were initially on anticoagulation continued their treatment (p<0.001).

**Table 1.** General characteristics of study population according to the treatment after HT.

|  | All (n = 763) | AC (n = 324) | APT (n = 191) | NM (n = 248) | p-value |
| --- | --- | --- | --- | --- | --- |
| Age | 74.6±8.9 | 74.5±9.0 | 74.5±8.7 | 74.9±9.0 | 0.824 |
| Male | 367 (48.1%) | 149 (46.0%) | 85 (44.5%) | 133 (53.6%) | 0.100 |
| HT Type |  |  |  |  | <.001 |
| HI-1 | 347 (45.5%) | 189 (58.3%) | 97 (50.8%) | 61 (24.6%) |  |
| HI-2 | 322 (42.2%) | 126 (38.9%) | 78 (40.8%) | 118 (47.6%) |  |
| PH-1 | 94 (12.3%) | 9 (2.8%) | 16 (8.4%) | 69 (27.8%) |  |
| initial NIHSS score | 11 (6 - 15) | 9 (3 - 13) | 12 (7 - 15) | 12 (8 - 15) | <.001 |
| previous mRS score | 0 (0 - 1) | 0 (0 - 1) | 0 (0 - 1) | 0 (0 - 0) | 0.405 |
| BMI | 23.3±3.1 | 23.2±3.1 | 23.0±2.9 | 23.7±3.3 | 0.056 |
| ASPECT DWI score | 7 (5 - 8) | 7 (6 - 8) | 7 (5 - 8) | 7 (5 - 8) | <.001 |
| Anterior (86.2%) | 7 (5 - 8) | 7 (6 - 8) | 7 (5 - 8) | 7 (5 - 8) | <.001 |
| Posterior (13.8%) | 8 (7 - 8) | 8 (7 - 8) | 8 (7 - 8) | 8 (7 - 9) | 0.524 |
| WBC | 8.6±3.1 | 8.1±3.0 | 9.2±3.3 | 9.0±3.1 | <.001 |
| Total cholesterol | 161.0±35.9 | 158.3±35.5 | 167.3±36.3 | 159.8±35.7 | 0.017 |
| BUN | 18.4±8.0 | 17.6±7.8 | 18.8±8.4 | 19.1±7.9 | 0.069 |
| Creatinine | 0.86±0.40 | 0.86±0.34 | 0.81±0.40 | 0.92±0.47 | 0.015 |
| Hemoglobin | 13.4±1.8 | 13.3±1.8 | 13.3±1.8 | 13.5±1.8 | 0.483 |
| Fasting glucose | 135.3±47.9 | 131.3±50.3 | 132.3±45.9 | 142.8±45.5 | 0.010 |
| Platelet count | 204.5±59.2 | 204.7±59.5 | 210.3±60.5 | 199.9±57.5 | 0.189 |
| LDL-C | 97.5±28.8 | 96.6±27.7 | 100.7±30.6 | 96.1±28.9 | 0.196 |
| history of TIA | 12 (1.6%) | 8 (2.5%) | 0 (0.0%) | 4 (1.6%) | 0.061 |
| history of stroke | 186 (24.4%) | 93 (28.7%) | 51 (26.7%) | 42 (16.9%) | 0.004 |
| history of PAD | 11 (1.4%) | 1 (0.3%) | 3 (1.6%) | 7 (2.8%) | 0.037 |
| history of CHD | 63 (8.3%) | 24 (7.4%) | 19 (9.9%) | 20 (8.1%) | 0.594 |
| history of HTN | 486 (63.7%) | 200 (61.7%) | 132 (69.1%) | 154 (62.1%) | 0.198 |
| history of DM | 203 (26.6%) | 85 (26.2%) | 43 (22.5%) | 75 (30.2%) | 0.188 |
| history of DL | 93 (12.2%) | 39 (12.0%) | 26 (13.6%) | 28 (11.3%) | 0.757 |
| Smoking |  |  |  |  | 0.774 |
| Current smoking | 72 (9.4%) | 34 (10.5%) | 18 (9.4%) | 20 (8.1%) |  |
| Ex-smoking | 65 (8.5%) | 27 (8.3%) | 19 (9.9%) | 19 (7.7%) |  |
| Never | 626 (82.0%) | 263 (81.2%) | 154 (80.6%) | 209 (84.3%) |  |
| history of AC | 165 (21.6%) | 97 (29.9%) | 29 (15.2%) | 39 (15.7%) | <.001 |
| history of statin | 119 (15.6%) | 56 (17.3%) | 31 (16.2%) | 32 (12.9%) | 0.345 |
| history of DM treat | 154 (20.2%) | 64 (19.8%) | 30 (15.7%) | 60 (24.2%) | 0.087 |
| history of HTN treat | 396 (51.9%) | 157 (48.5%) | 109 (57.1%) | 130 (52.4%) | 0.165 |
| Acute treatment |  |  |  |  | 0.009 |
| IVT only | 157 (20.6%) | 45 (13.9%) | 51 (26.7%) | 61 (24.6%) |  |
| IAT only | 115 (15.1%) | 56 (17.3%) | 28 (14.7%) | 31 (12.5%) |  |
| IVT + IAT | 93 (12.2%) | 40 (12.3%) | 22 (11.5%) | 31 (12.5%) |  |
| None | 398 (52.2%) | 183 (56.5%) | 90 (47.1%) | 125 (50.4%) |  |
| Discharge APT | 259 (33.9%) | 72 (22.2%) | 123 (64.4%) | 64 (25.8%) | <.001 |
| Discharge AC | 396 (51.9%) | 267 (82.4%) | 56 (29.3%) | 73 (29.4%) | <.001 |
| mean SBP | 135.6±14.4 | 134.6±14.3 | 137.3±14.2 | 135.5±14.5 | 0.117 |

† P-value by Chi-square test, Fisher's exact test, Cochran-Mantel-Haenszel shift test, ANOVA and Kruskal-Wallis Test
AC, anticoagulation; APT, antiplatelet; NM, no or quit medication; HT, hemorrhagic transformation; HI, hemorrhagic infarction; PH, parenchymal hematoma; NIHSS, National institute of health stroke scale; BMI, body mass index; WBC, white blood cell; BUN, blood urea nitrogen; LDL-C, low density lipoprotein; TIA, transient cerebral ischemia; PAD, peripheral arterial disease; CHD, coronary heart disease; HTN, hypertension; DM, diabetes mellitus; DL, dyslipidemia; IVT, intravenous thrombolysis; IAT, intra-arterial thrombectomy; SBP, systolic blood pressure

### Primary outcome: HT exacerbation and mRS shift

HT exacerbation was observed in 28.7% of patients based on imaging findings, with rates of 30.9% in the AC group, 21.5% in the APT group, and 31.5% in the NM group (Table 2). In the unadjusted model, HT exacerbation was more common in the AC group compared to the APT group (unadjusted OR 0.61, 95% CI 0.40-0.93, p=0.022), while there was no significant difference compared to the NM group (unadjusted OR 1.03, 95% CI 0.72-1.47, p=0.880). The shift towards a better outcome in mRS scores at 3 months was more likely to be achieved in the AC group compared to the NM group (unadjusted OR 0.26, 95% CI 0.19-0.36, p<0.001). These trends persisted even after adjusting for variables. The use of AC was associated with higher HT exacerbation compared to the APT group (aOR 0.48, 95% CI 0.29-0.80, p=0.005), while there was no statistically significant difference compared to the NM group (aOR 0.86, 95% CI 0.53-1.37, p=0.521). Additionally, the mRS shift at 3 months indicated better outcomes with AC compared to both APT (aOR 0.63, 95% CI 0.43-0.92, p=0.017) and NM (aOR 0.38, 95% CI 0.26-0.55, p<0.001) (Table 3). The mRS distribution at 3 months is presented in Figure 1, showing significantly better outcomes in the AC group compared to the APT and NM groups.

**Figure 1.**
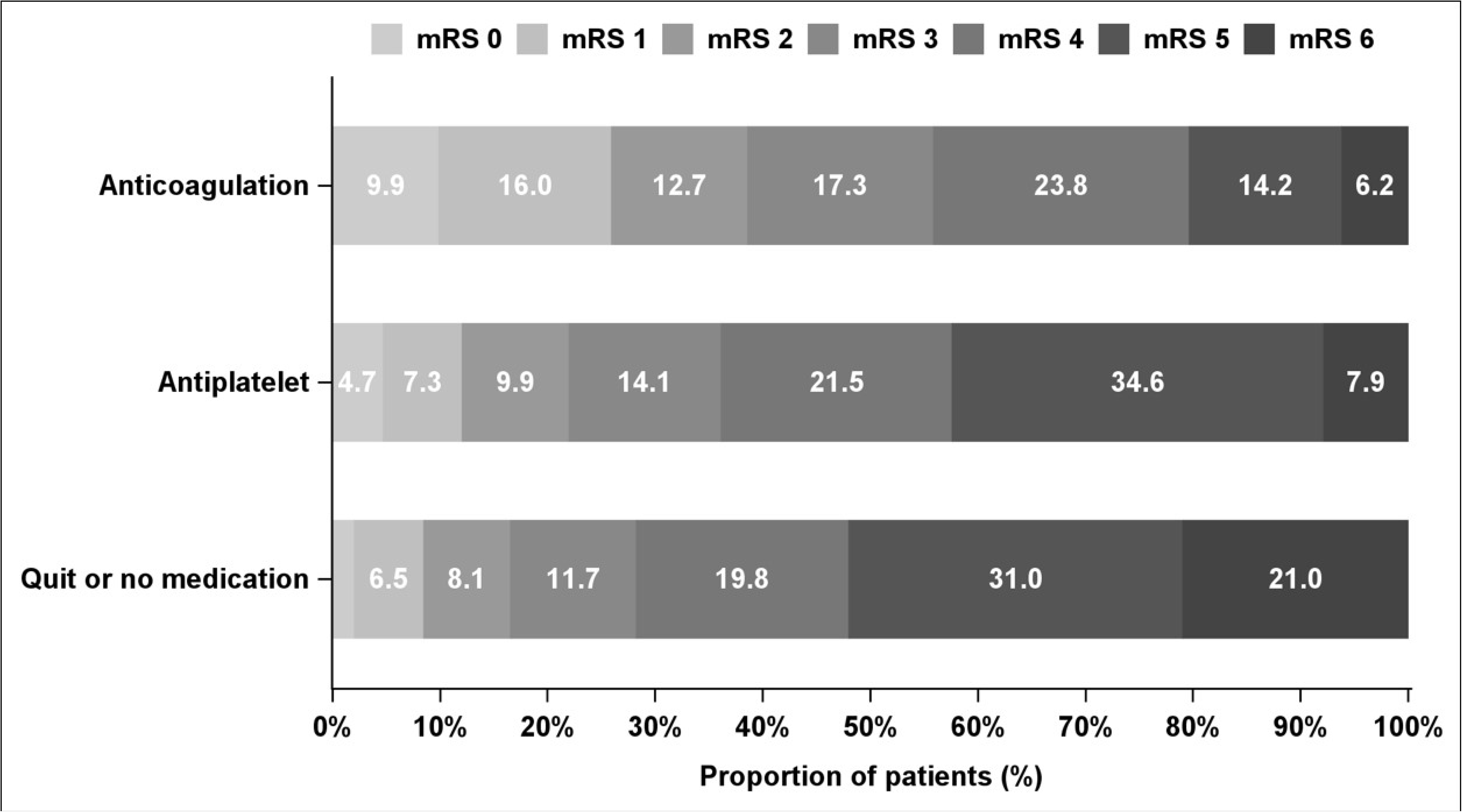
3-month mRS distribution according to the treatment after HT

**Table 2.** Event rates of outcomes according to the treatment after HT.

| Event Rate | All | AC | APT | NM | p-value |
| --- | --- | --- | --- | --- | --- |
| **Number** | 763 | 324 | 191 | 248 |  |
| HT exacerbation in imaging |  |  |  |  | 0.038 |
| Number of events | 219 | 100 | 41 | 78 |  |
| Event rates (% , 95% CI) | 28.7 (25.5-32.1) | 30.9 (25.9-36.2) | 21.5 (15.9-28.0) | 31.5 (25.7-37.6) |  |
| 3 months good mRS (0-2) |  |  |  |  | <.001 |
| Number of events | 208 | 125 | 42 | 41 |  |
| Event rates (% , 95% CI) | 27.3 (24.1-30.6) | 38.6 (33.3-44.1) | 22.0 (16.3-28.5) | 16.5 (12.1-21.8) |  |
| Symptomatic ICH |  |  |  |  | <.001 |
| Number of events | 38 | 5 | 4 | 29 |  |
| Event rates (% , 95% CI) | 5.0 (3.5-6.8) | 1.5 (0.5-3.6) | 2.1 (0.6-5.3) | 11.7 (8.0-16.4) |  |
| Recurrence of stroke in 3-month |  |  |  |  | 0.047 |
| Number of events | 18 | 5 | 9 | 4 |  |
| Event rates (% , 95% CI) | 2.4 (1.3-3.4) | 1.5 (0.2-2.9) | 4.7 (1.7-7.7) | 1.6 (0.0-3.2) |  |
| Composite vascular outcome in 3-month |  |  |  |  | <.001 |
| Number of events | 95 | 24 | 22 | 49 |  |
| Event rates (% , 95% CI) | 12.5 (10.1-14.8) | 7.4 (4.6-10.3) | 11.5 (7.0-16.0) | 19.8 (14.8-24.7) |  |

P-value by Chi-square test and log rank test
APT, antiplatelet; AC, anticoagulation; NM, no or quit medication; HT, hemorrhagic transformation; mRS, modified Rankin Scale; ICH, intracerebral hemorrhage

**Table 3.**
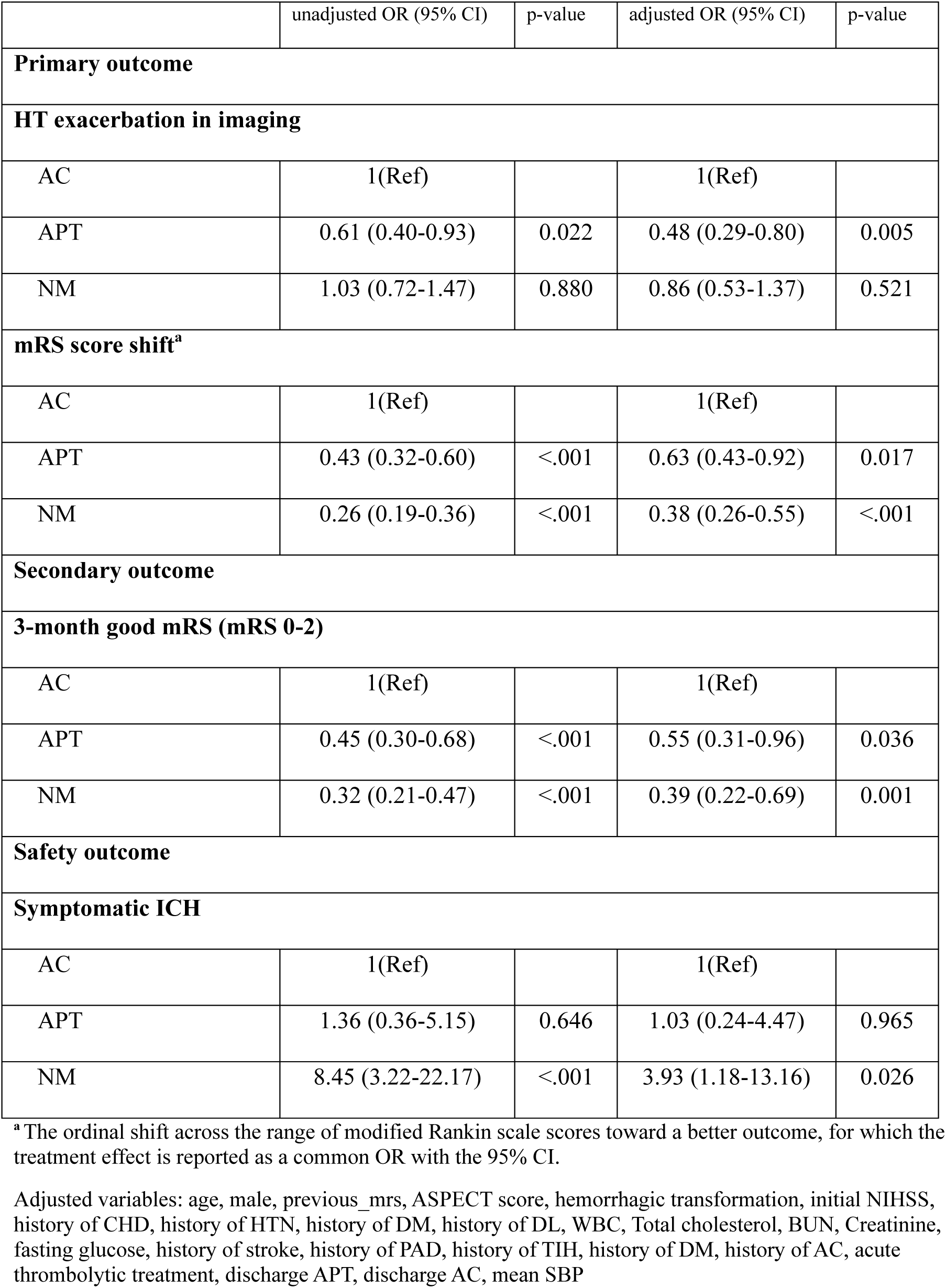
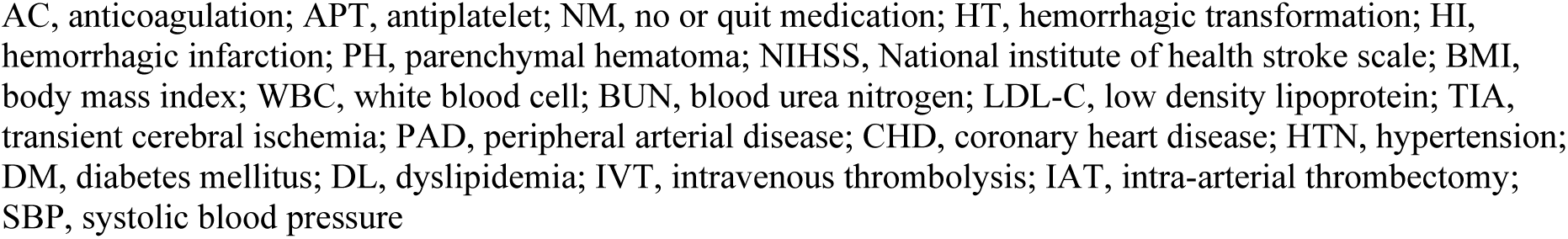
Associations of the treatment after HT with outcomes.

|  | unadjusted OR (95% CI) | p-value | adjusted OR (95% CI) | p-value |
| --- | --- | --- | --- | --- |
| **Primary outcome** |  |  |  |  |
| **HT exacerbation in imaging** |  |  |  |  |
| AC | 1(Ref) |  | 1(Ref) |  |
| APT | 0.61 (0.40-0.93) | 0.022 | 0.48 (0.29-0.80) | 0.005 |
| NM | 1.03 (0.72-1.47) | 0.880 | 0.86 (0.53-1.37) | 0.521 |
| **mRS score shift<sup>a</sup>** |  |  |  |  |
| AC | 1(Ref) |  | 1(Ref) |  |
| APT | 0.43 (0.32-0.60) | <.001 | 0.63 (0.43-0.92) | 0.017 |
| NM | 0.26 (0.19-0.36) | <.001 | 0.38 (0.26-0.55) | <.001 |
| **Secondary outcome** |  |  |  |  |
| **3-month good mRS (mRS 0-2)** |  |  |  |  |
| AC | 1(Ref) |  | 1(Ref) |  |
| APT | 0.45 (0.30-0.68) | <.001 | 0.55 (0.31-0.96) | 0.036 |
| NM | 0.32 (0.21-0.47) | <.001 | 0.39 (0.22-0.69) | 0.001 |
| **Safety outcome** |  |  |  |  |
| **Symptomatic ICH** |  |  |  |  |
| AC | 1(Ref) |  | 1(Ref) |  |
| APT | 1.36 (0.36-5.15) | 0.646 | 1.03 (0.24-4.47) | 0.965 |
| NM | 8.45 (3.22-22.17) | <.001 | 3.93 (1.18-13.16) | 0.026 |

<sup>a</sup> The ordinal shift across the range of modified Rankin scale scores toward a better outcome, for which the treatment effect is reported as a common OR with the 95% CI.
Adjusted variables: age, male, previous\_mrs, ASPECT score, hemorrhagic transformation, initial NIHSS, history of CHD, history of HTN, history of DM, history of DL, WBC, Total cholesterol, BUN, Creatinine, fasting glucose, history of stroke, history of PAD, history of TIH, history of DM, history of AC, acute thrombolytic treatment, discharge APT, discharge AC, mean SBP
AC, anticoagulation; APT, antiplatelet; NM, no or quit medication; HT, hemorrhagic transformation; HI, hemorrhagic infarction; PH, parenchymal hematoma; NIHSS, National institute of health stroke scale; BMI, body mass index; WBC, white blood cell; BUN, blood urea nitrogen; LDL-C, low density lipoprotein; TIA, transient cerebral ischemia; PAD, peripheral arterial disease; CHD, coronary heart disease; HTN, hypertension; DM, diabetes mellitus; DL, dyslipidemia; IVT, intravenous thrombolysis; IAT, intra-arterial thrombectomy; SBP, systolic blood pressure

**Table 4.** Association of the treatment after HT with vascular outcomes.

|  | unadjusted HR (95% CI) | p-value | adjusted HR (95% CI) | p-value |
| --- | --- | --- | --- | --- |
| **Secondary outcome** |  |  |  |  |
| **3-month stroke recurrence** |  |  |  |  |
| AC | 1(Ref) |  | 1(Ref) |  |
| APT | 3.09 (1.04-9.23) | 0.043 | 2.86 (0.75-10.91) | 0.124 |
| NM | 1.05 (0.28-3.91) | 0.942 | 1.38 (0.30-6.24) | 0.677 |
| **3-month composite vascular outcome** |  |  |  |  |
| AC | 1(Ref) |  | 1(Ref) |  |
| APT | 1.59 (0.89-2.83) | 0.118 | 1.25 (0.64-2.45) | 0.518 |
| NM | 2.88 (1.77-4.69) | <.001 | 2.01 (1.09-3.72) | 0.025 |

Adjusted variables: age, male, previous\_mrs, ASPECT score, hemorrhagic transformation, initial NIHSS, history of CHD, history of HTN, history of DM, history of DL, WBC, Total cholesterol, BUN, Creatinine, fasting glucose, history of stroke, history of PAD, history of TIH, history of DM, history of AC, acute thrombolytic treatment, discharge APT, discharge AC, mean SBP
AC, anticoagulation; APT, antiplatelet; NM, no or quit medication; HT, hemorrhagic transformation; HI, hemorrhagic infarction; PH, parenchymal hematoma; NIHSS, National institute of health stroke scale; BMI, body mass index; WBC, white blood cell; BUN, blood urea nitrogen; LDL-C, low density lipoprotein; TIA, transient cerebral ischemia; PAD, peripheral arterial disease; CHD, coronary heart disease; HTN, hypertension; DM, diabetes mellitus; DL, dyslipidemia; IVT, intravenous thrombolysis; IAT, intra-arterial thrombectomy; SBP, systolic blood pressure

### Safety outcome and secondary outcomes

sICH occurred in 5% of cases overall, with incidence rates of 1.5% in the AC group, 2.1% in the APT group, and 11.7% in the NM group (p <0.001) (Table 2). In the unadjusted model, the NM group had 8.45 times higher sICH risk compared to the AC group, and this trend persisted even after adjusting for variables (aOR 3.93, 95% CI 1.18-13.16, p=0.026) (Table 3).

At 3 months, the proportion of patients 3-month mRS 0-2, considered a good functional outcome, was 38.6% in the AC group compared to 16.5% in the NM group (Table 2). In crude analysis, the AC group had a higher rate of good functional outcomes compared to other treatments. Even after adjusting for variables, the likelihood of achieving mRS 0-2 at 3 months was significantly higher, with a 45% greater chance compared to the APT group and a 61% greater chance compared to the NM group (Table 3). At 3 months, the composite vascular outcome, including stroke, myocardial infarction (MI), and mortality, occurred in 12.5% of all cases, with statistical differences observed at 7.4% in the AC group, 11.5% in the APT group, and 19.8% in the NM group (p<0.001). Stroke recurrence at 3 months occurred in 2.4% of patients overall, with rates of 1.5% in the AC group, 4.7% in the APT group, and 1.6% in the NM group, showing differences in incidence between groups (p=0.047) (Table 2). In the unadjusted model, the 3-month composite outcome was 2.88 times higher in the NM group compared to the AC group, and this significance remained after adjusting for variables (adjusted HR 2.01, 95% CI 1.09-3.72, p=0.025). Stroke recurrence showed a higher risk in the APT group compared to the AC group in the unadjusted model (unadjusted HR 3.09, 95% CI 1.04-9.23, p=0.043), although this significance was not maintained after adjusting for variables (adjusted HR 2.86, 95% CI 0.75-10.91, p=0.124).

The Kaplan-Meier survival curve for these outcomes is presented in Figure 2, demonstrating differences in stroke recurrence and composite vascular outcomes across treatment groups.

**Figure 2.**
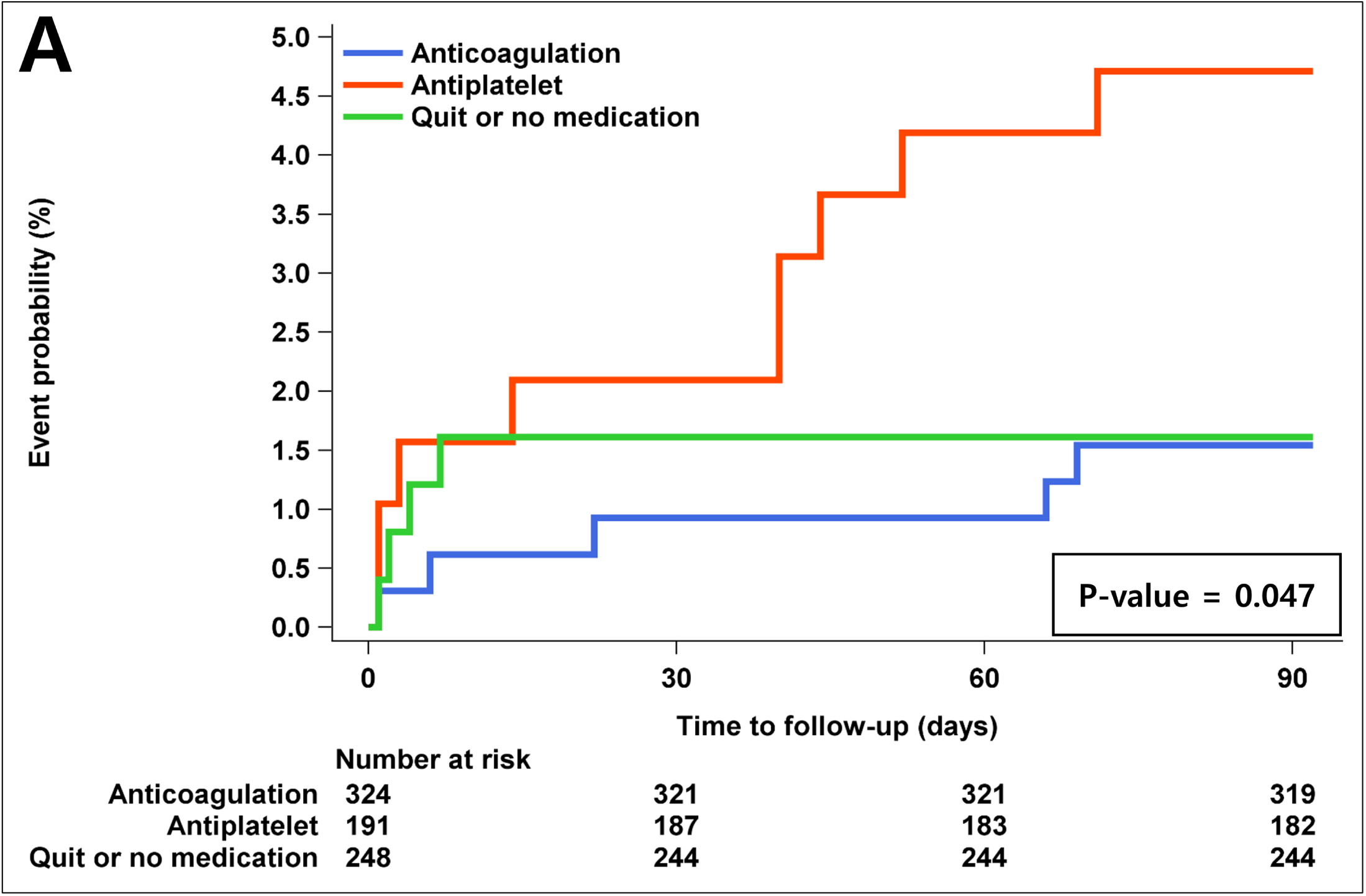
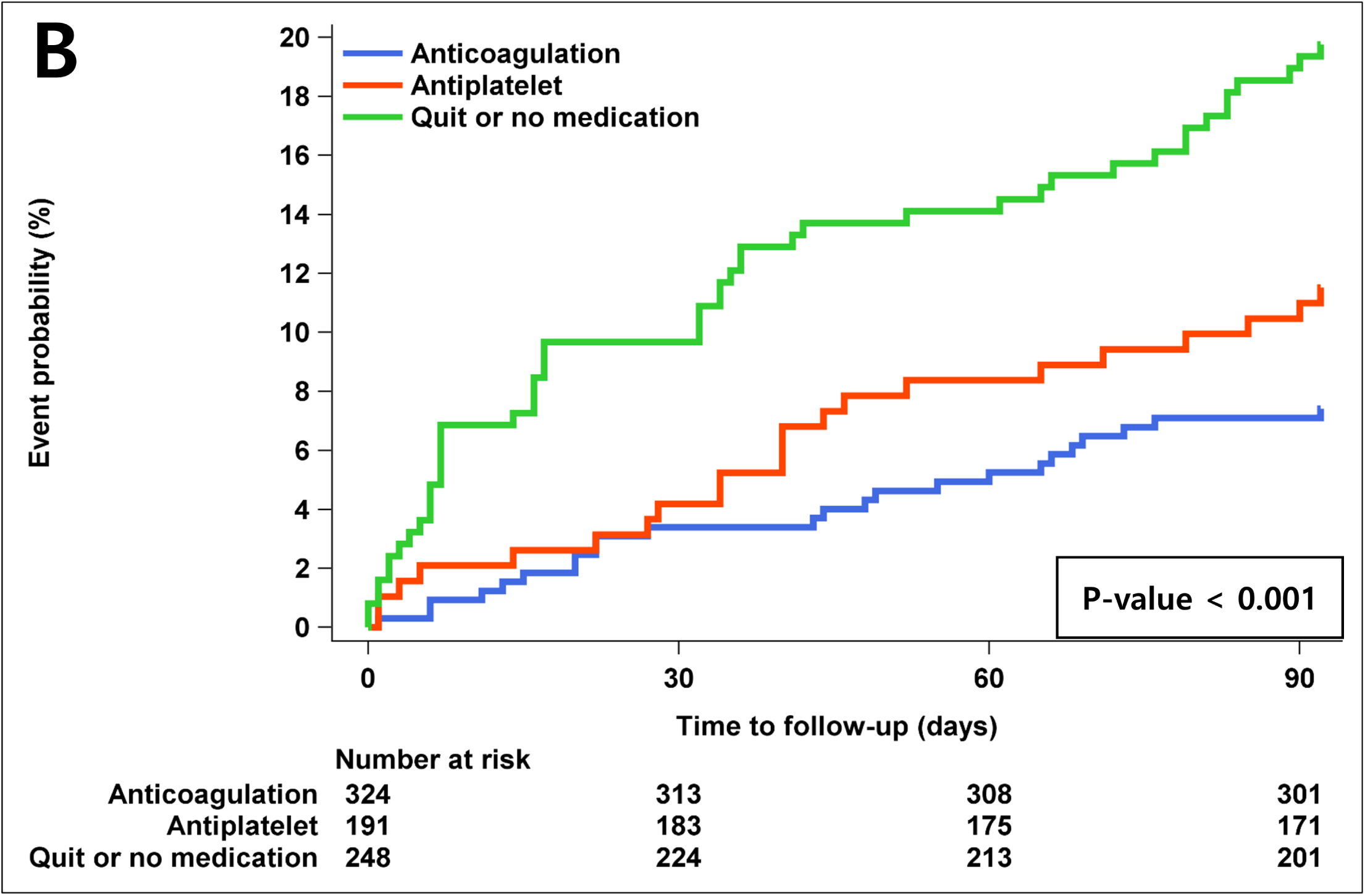
Associations of the treatment after HT with 3-month vascular outcomes. A) Stroke recurrence, B) composite vascular outcome

## Discussion

Our investigation sought to elucidate the radiological and clinical implications of anticoagulation therapy in patients with acute cardioembolic stroke developed HT. While our findings suggest that anticoagulation may be associated with radiological exacerbation of HT compared to other treatment modalities, this did not correlate with an increased incidence of sICH. Notably, patients receiving anticoagulation during HT exhibited significantly improved functional outcomes at the 3-month follow-up. This observation underscores the potential benefits of anticoagulation in facilitating long-term recovery, despite prevailing concerns regarding HT progression.

The heightened prevalence of HT in cardioembolic stroke can be attributed to the propensity for larger infarct volumes and compromised collateral circulation, both of which contribute to an elevated risk of HT. Furthermore, the incidence of PH is demonstrably higher in cardioembolic stroke compared to other etiologies.^20, 21^ Additionally, the use of anticoagulant therapy for secondary prevention further increases the risk of HT.^22–24^ Despite concerns that anticoagulation could exacerbate HT, our results indicate that careful patient selection and appropriate timing of anticoagulation initiation can mitigate this risk, leading to improved clinical outcomes.

The precise mechanism by which anticoagulation affects HT following its onset remains elusive. HT primarily results from blood-brain barrier disruption due to ischemic damage, often exacerbated by inflammatory and proteolytic cascades, including the activation of matrix metalloproteinases and increased oxidative stress. Interestingly, while anticoagulation therapies effectively mitigate thromboembolic risks by inhibiting new clot formation, they do not necessarily amplify these proteolytic pathways. Therefore, their effect on exacerbating HT, especially in non-massive forms like HI-1 or HI-2, is limited. Current research suggests that the etiology of HT is multifactorial, extending beyond the sole influence of anticoagulation. The judicious management of systemic and neuroinflammatory factors may play a crucial role in mitigating the risk of further bleeding. This complex interplay underscores the need for a nuanced approach to anticoagulation in the context of HT, balancing the prevention of further ischemic events against the management of potential hemorrhagic complications.^6, 25, 26^

Our findings corroborate the results of several recent large-scale trials that advocate for early anticoagulation in patients with atrial fibrillation and AIS. Notably, studies such as the ELAN and TIMING trials have demonstrated that early initiation of anticoagulation is non-inferior to delayed initiation in terms of both safety and efficacy, particularly in preventing recurrent ischemic events.^13, 14^ Consistent with our observations, these trials reported improved functional outcomes associated with early anticoagulation without a significant increase in sICH.

A subgroup analysis of the ELAN trial,^15^ which evaluated the potential benefits of initiating direct oral anticoagulants (DOACs) earlier than guideline recommendations in AIS patients, examined outcomes based on the presence or absence of HT. The results showed no significant difference between early and late initiation of DOACs in terms of efficacy and safety, regardless of HT status. For patients with HT, the adjusted risk difference for early treatment compared to late treatment was -2.2% (95% CI -7.8% to 3.5%), with no difference in the incidence of sICH, suggesting that early OAC in the presence of HT does not significantly compromise safety and may even be beneficial. However, when focusing specifically on patients with PH, early OAC was associated with a 25.1% higher likelihood of a poor 3-month outcome (mRS 3-6). Our study design differs slightly, as it investigates drug use when HT is already present rather than at the time of drug initiation, making it distinct from the subgroup analysis of ELAN trial. Nonetheless, both studies indicate that OAC can be used relatively safely even in the presence of HT and may positively influence functional prognosis.

Another study analyzed the worsening of HT in relation to warfarin use compared to no medication and antiplatelet therapy among 220 patients who developed HT at a single stroke center in a tertiary hospital.^16^ The study reported HT aggravation in 14.3% of the warfarin group and 4.2% in the antiplatelet group, although no statistically significant difference was observed between the two cohorts. However, this study’s limitations include its lack of focus on cardioembolic stroke exclusively and the relatively small sample size of warfarin users (n=28), which constrains the clarity of its clinical significance. In contrast, our investigation analyzed the relationship between HT and medication use specifically in a cohort of approximately 700 patients with cardioembolic stroke, providing a more robust and targeted analysis.

At our center, calculating infarct size during image analysis was not available. Since most of our centers used DWI as the initial examination, we attempted to adjust for severity using the ASPECT score derived from DWI images. Malignant cerebral infarctions with an ASPECT score between 0 and 2, which were anticipated to have a poor prognosis and thus were not considered candidates for active treatment, were excluded from the analysis. Additionally, the ASPECT score was measured separately for the anterior circulation and posterior circulation.

However, due to the small proportion of cases involving the posterior circulation, we were unable to analyze differences in outcomes based on stroke location.

Our findings indicate that APT use in the presence of HT was associated with 52% less radiological deterioration compared to AC use and showed no significant difference compared to NM cases. Notably, continuing AC did not increase the likelihood of sICH compared to other treatments. In fact, the NM group exhibited a 3.93-fold higher incidence of sICH. These results suggest that HT progression to sICH in AIS may be part of the natural course of stroke, potentially more related to the initial HT type rather than medication use. This implies that if certain initial HT types can be avoided, early AC could potentially improve stroke prognosis. Nevertheless, better-designed studies are needed in the future to validate these findings. Additionally, in our study, the AC group tended to have lower initial severity and higher ASPECT scores. While these factors could potentially influence functional outcomes, we adjusted for both initial severity and ASPECT scores in our analysis to account for this potential confounding, ultimately yielding positive results.

Several limitations of our study warrant consideration. Firstly, despite analyzing a large number of HT patients, the single-center design introduces potential center bias, limiting generalizability. Secondly, the retrospective nature of the study presents inherent limitations. Thirdly, image analysis relied on individual expert judgment, which may introduce variability in interpretation. We aimed to mitigate this by conducting regular meetings to review and standardize analyses. Fourthly, our classification of treatment methods did not distinguish between different drug doses or types, necessitating further research with larger cohorts.

Despite these limitations, our study remains significant as a relatively large-scale investigation focused exclusively on patients who developed HT.

In conclusion, our study suggests that AC administration in acute cardioembolic stroke with HT, excluding PH-2, may lead to radiological worsening but does not result in increased sICH and is associated with improved functional outcomes. The presence of HT alone should not necessarily preclude AC therapy. The outcomes of drug administration following HT may vary depending on the initial HT type. Further robust, large-scale prospective studies are essential to validate these findings.

## Data Availability

The data used in this study are available upon reasonable request to corresponding author.

## ADDITIONAL INFORMATION

### Declaration of Competing interests

The authors declare that they have no competing interests.

### Disclosure

None

### Funding

This study was supported by a grant (BCRI24011) of Chonnam National University Hospital Biomedical Research Institute.

### Data availability

Data used in this study are available upon reasonable request to corresponding author.

## Acknowledgements

None

## Authors’ contributions

- Study concept and design: JT Kim, H Kim
- Acquisition of data: H Kim, JM Kim, JT Kim, MS Park, KH Choi, BS Seo, YE An
- Analysis and interpretation of data: JS Lee
- Drafting of the manuscript: JT Kim, H Kim
- All authors read and approved the final manuscript.

## Notes

### Competing Interest Statement

The authors have declared no competing interest.

### Funding Statement

.No external funding was received for this study.

### Author Declarations

The current study was approved by local institutional review boards at all participating centers, including Chonnam National University Hospital (CNUH-2024-294).

